# COVID-19 early warning score: a multi-parameter screening tool to identify highly suspected patients

**DOI:** 10.1101/2020.03.05.20031906

**Authors:** Cong-Ying Song, Jia Xu, Jian-Qin He, Yuan-Qiang Lu

## Abstract

**BACKGROUND:** Corona Virus Disease 2019 (COVID-19) is spreading worldwide. Effective screening for patients is important to limit the epidemic. However, some defects make the currently applied diagnosis methods are still not very ideal for early warning of patients. We aimed to develop a diagnostic model that allows for the quick screening of highly suspected patients using easy-to-get variables.

**METHODS:** A total of 1,311 patients receiving severe acute respiratory syndrome coronavirus 2 (SARS-CoV-2) nucleicacid detection were included, whom with a positive result were classified into COVID-19 group. Multivariate logistic regression analyses were performed to construct the diagnostic model. Receiver operating characteristic (ROC) curve analysis were used for model validation.

**RESULTS:** After analysis, signs of pneumonia on CT, history of close contact, fever, neutrophil-to-lymphocyte ratio (NLR), Tmax and sex were included in the diagnostic model. Age and meaningful respiratory symptoms were enrolled into COVID-19 early warning score (COVID-19 EWS). The areas under the ROC curve (AUROC) indicated that both of the diagnostic model (training dataset 0.956 [95%CI 0.935-0.977, *P* < 0.001]; validation dataset 0.960 [95%CI 0.919-1.0, *P* < 0.001]) and COVID-19 EWS (training dataset 0.956 [95%CI 0.934-0.978, *P* < 0.001]; validate dataset 0.966 [95%CI 0.929-1, *P* < 0.001]) had good discrimination capacity. In addition, we also obtained the cut-off values of disease severity predictors, such as CT score, CD8^+^ T cell count, CD4^+^ T cell count, and so on.

**CONCLUSIONS:** The new developed COVID-19 EWS was a considerable tool for early and relatively accurately warning of SARS-CoV-2 infected patients.

## INTRODUCTION

The Corona Virus Disease 2019 (COVID-19), caused by severe acute respiratory syndrome coronavirus 2 (SARS-CoV-2) infection is erupting worldwide.^1-4^ Due to the negative effects it brought, the World Health Organization (WTO) defined the outbreak a public health emergency of international concern on January 31th, 2020.^5^ As early as December 2019, the COVID-19 epidemic were reported in Wuhan, Hubei Province, China. In a short period of time, the number of COVID-19 confirmed patients in Zhejiang Province also increase rapidly.^6,7^

Currently, the diagnosis of COVID-19 relies mainly on SARS-CoV-2 nucleicacid detection. However, due to the shortcomings of false-negative results caused by the low viral load in the samples and relatively insufficient detection kits, many patients cannot be detected in time. Although clinical symptoms, epidemiological exposure history and computed tomography (CT) manifestations have been added to the diagnostic criteria of the disease, the atypical early clinical symptoms, unclear epidemiological exposure history and ambiguous imaging conclusions also bring difficulties to the screening of the patients.^6,8^ An effective and simple multi-parameter diagnostic tool is urgently needed.

Our study selected the clinical data of patients who came to our hospital during the first phase of the epidemic (by February 5^th^, 2020, most of the confirmed patients were from Wuhan or had a clear history of close contact with the returning population from Wuhan) as the training dataset. By comparing the characteristics of the confirmed cases and excluding cases, the independent risk factors related to the disease were identified and included in the diagnostic model. The purpose of establishing a multifaceted diagnostic model is to improve the detection rate and detection speed of infected patients, so as to truly achieve early detection, early isolation and early treatment to curb the further development of the epidemic.

## METHODS

### POPULATIONS

We included clinical data of a total of 1,311 patients who came to the First Affiliated Hospital, School of Medicine, Zhejiang University and performed at least once SARS-CoV-2 nucleicacid detection for analysis. This study was approved by the Ethical Committee of the First Affiliated Hospital, School of Medicine, Zhejiang University (code number IIT20200025A). Data of patients between January 20^th^, 2020 and February 5^th^, 2020 were enrolled into training dataset, and between February 6^th^, 2020 and February 19^th^, 2020 were enrolled into validate dataset. All patients who had a positive SARS-CoV-2 nucleicacid detection result were classified into COVID-19 group, while the others were classified into non-COVID-19 group. Due to the huge number of non-COVID-19 group patients, we eliminated the enrolled patients according to the following steps: (1) all negative patients from the COVID-19 designated district, Zhijiang District of the First Affiliated Hospital of Zhejiang University School of Medicine; (2) negative outpatients from Qingchun District were sorted by “Z-A” based on Chinese first name, the first 60% between January 20^th^, 2020 and February 5^th^, 2020, and the first 40% between February 6^th^, 2020 and February 19^th^, 2020 were included. Exclusion criteria: (1) asymptomatic patients without history of exposure but had strong willingness for detection; (2) patients with important information deficits. Figure 1 presented the concrete procedures of patients screening. The definitions of severity was based on the New Coronavirus Pneumonia Prevention and Control Program (6th edition) published by the National Health Commission of China^9^: (1) mild: patients had no pneumonia on imaging;(2) moderate: patients with symptoms and imaging examination shows pneumonia; (3) severe: patients meet any of the following: (i) respiratory rate ≥ 30/min; (ii) resting pulse oxygen saturation (SpO_2_) ≤ 93%; (iii) partial pressure of oxygen (PaO_2_) / fraction of inspired oxygen (FiO_2_) ≤ 300mmHg (1mmHg = 0.133kPa); (iiii) multiple pulmonary lobes showing more than 50% progression of lesion in 24-48 hours on imaging; (4) critical: patients meet any of the following: (i) respiratory failure requires mechanical ventilation; (ii) shock; (iii) combination of other organ failure that requires admitted into intensive care unit (ICU). Mild or moderate patients were included in the non-severe group, and severe or critical patients were included in the severe group.

**Figure 1.**
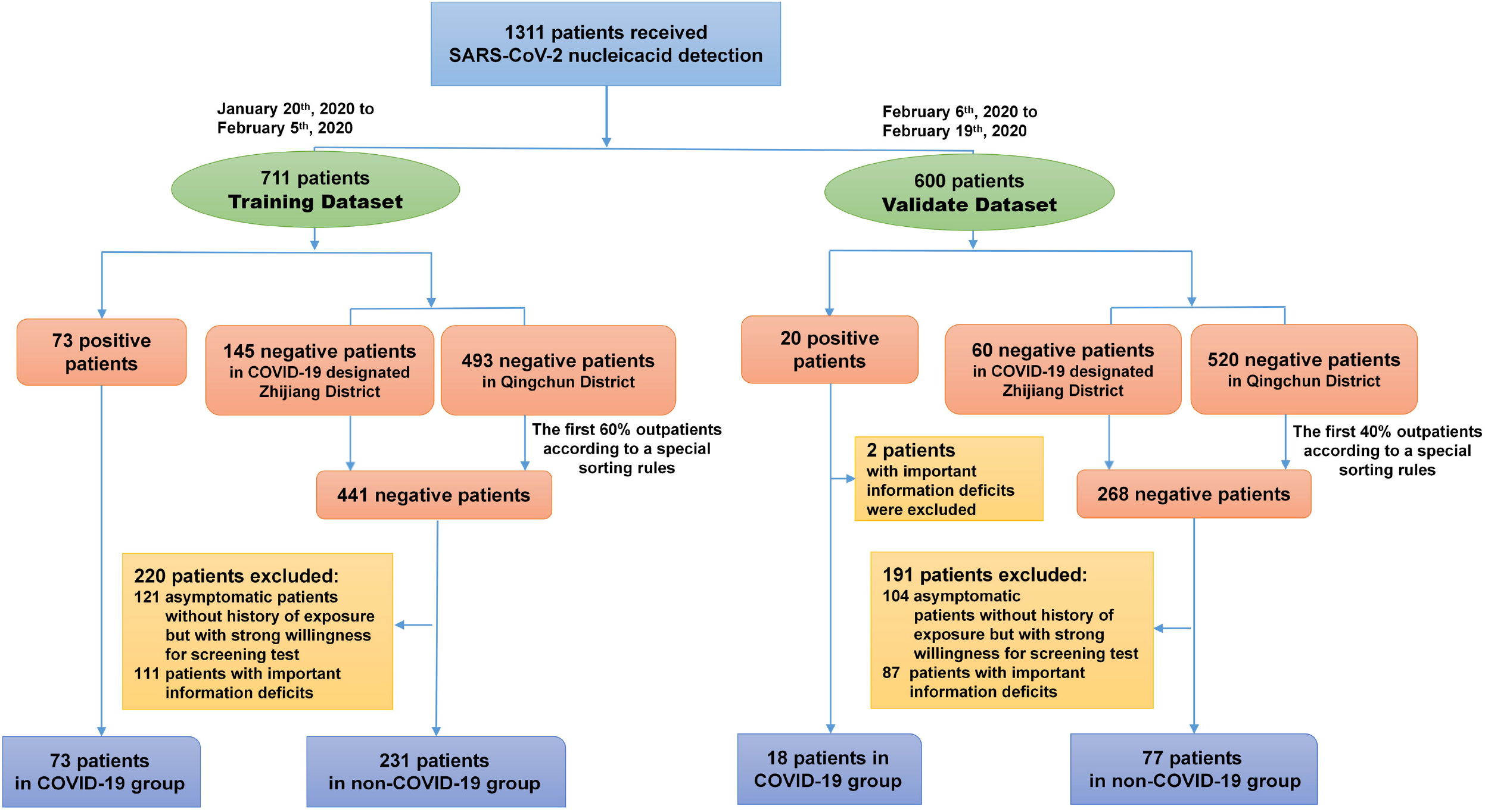
Flow chart of patients screening. Shown were the concrete procedures of patients screening. A total of 1,311 patients received SARS-CoV-2 nucleicacid detection in our hospital were enrolled. According to the time distribution, these patients were divided into the training dataset and the validate dataset. Base on the results of SARS-CoV-2 nucleicacid detection, patients were enrolled into COVID-19 group or non-COVID-19 group. The whole screening process strictly followed the inclusion criteria and exclusion criteria.

### DATA COLLECTION

Data were extracted from electronic medical record system of the the First Affiliated Hospital, School of Medicine, Zhejiang University. Epidemiological exposure history within the 14 days before illness onset is defined as: (1) having exposure related to Wuhan: had recently lived, traveled, or had close contact with someone who has been to Wuhan; (2) close contacts: had contacted with a COVID-19 confirmed patient. Except for SARS-CoV-2 nucleicacid detection, the data used in this study were the first inhospital results. Based on the clinical judgment of doctors and the medical burden considerations, patients who did not performed CT in non-COVID-19 group were assigned as a negative CT report by default.

### LABORATORY CONFIRMATION

Laboratory confirmation was achieved by real time reverse transcription-polymerase chain reaction (RT-PCR) assay for SARS-CoV-2 according to the protocol established by the WHO.^10^ Sputum samples were preferred in our hospital for RT-PCR assay within 3 hours. Two target genes of SARS-CoV-2 were tested during the process, including open reading frame 1ab (ORF1ab) and nucleocapsidprotein (N).

### CT SCORING

The CT scores were analyzed retrospectively by two radiologists blinded to the patient’s diagnosis and other clinical features. Two lung is divided into the upper (above trachea juga), middle (between trachea juga and pulmonary vein) and lower (under pulmonary vein) parts. According to the area occupied by different CT signs in each region: normal lung tissue, 0 points; lesion area < 25% of this layer, 1 point; 25% ∼ 50%, 2 points; 50% ∼ 75%, 3 points; > 75% 4 points. The score of each part were added to obtain the final CT score.^11^

### STATISTICAL ANALYSIS

Continuous variables were expressed as medians with interquartile ranges (IQR). Categorical variables were expressed as numbers and percentages in each category. Mann-Whitney U-test were used to evaluate continuous data. Chi-square test were used for categorical variables. Independent risk factor analysis was performed using multivariate logistic regression analyses with forward stepwise method. The discrimination capacity of the diagnostic model and COVID-19 early warning score (COVID-19 EWS) were accessed by calculating the area under the receiver operating characteristic (AUROC) curve. All statistical analyses were done by SPSS statistical software package (version 25.0). *P* < 0.05 was considered statistically significant.

## RESULTS

### DEMOGRAPHIC AND CLINICAL CHARACTERISTICS OF PATIENTS IN THE TRAINING DATASET

A total of 304 patients were enrolled in the training dataset, including 73 patients in the COVID-19 group and 231 patients in the non-COVID-19 group (Table 1). There was a higher proportion of males in the COVID-19 group (63.0% vs 37.0%, *P* = 0.007). Patients in COVID-19 group had a higher age distribution than non-COVID-19 group (53 years [IQR, 43.5-62.0] vs 34 years [IQR, 29.0-49.0]; *P* < 0.001). As for epidemiological exposure, proportion of the patients with history of Wuhan-related exposure and close contact was higher than the COVID-19 group (57.5% vs 40.7%, P = 0.012; 34.2% vs 5.6%, *P* < 0.001). Patients in the COVID-19 group had a longer out-hospital disease course than the non-COVID-19 group (7 [IQR, 4.0-9.0] vs 3 [IQR, 1.0-5.0]; *P* < 0.001). More patients in the COVID-19 group showed fever (94.5% vs 68.0%, *P* < 0.001), dyspnea (21.9% vs 8.7%, *P*= 0.002), cough (60.3% vs 45.9%, *P* = 0.032), expectoration (23.3% vs 12.6%, *P* = 0.026), and myalgia or fatigue (32.9% vs 19.9%, *P* = 0.022). On the other hand, more patients in the non-COVID-19 group showed pharyngalgia (22.9% vs 6.8%, *P* = 0.002), and rhinobyon or rhinorrhoea (10.4% vs 1.4%, *P* = 0.014). In addition, patients in the COVID-19 group had a higher maximum body temperature (Tmax) during the outhospital phase (38.2 °c [IQR,37.5-39.0], *P* < 0.001) and a lower SpO_2_ (97.0% [IQR, 95.0-99.0] vs 98.0% [98.0-99.0], *P* < 0.001).

**Table 1.** Clinical characteristics of patients in the training dataset.

| Demographic and clinical characteristics | All patients<br>(n = 304) | Non-COVID-19 patients<br>(n = 231) | COVID-19 patients<br>(n = 73) | P value |
| --- | --- | --- | --- | --- |
| **Age (years)** | 37.5 (30.0-55.0) | 34.0 (29.0-49.0) | 53.0 (43.5-62.0) | < 0.001 |
| **Sex** |  |  |  | 0.007 |
| Male | 150 (49.3) | 104 (45.0) | 46 (63.0) |  |
| Female | 154 (50.7) | 127 (55.0) | 27 (37.0) |  |
| **Symptoms** |  |  |  |  |
| Fever | 226 (74.3) | 157 (68.0) | 69 (94.50) | < 0.001 |
| Cough | 150 (49.3) | 106 (45.9) | 44 (60.3) | 0.032 |
| Expectoration | 46 (15.1) | 29 (12.6) | 17 (23.3) | 0.026 |
| Headache | 43 (14.1) | 34 (14.7) | 9 (12.3) | 0.610 |
| Myalgia or fatigue | 70 (23.0) | 46 (19.9) | 24 (32.9) | 0.022 |
| Chill | 26 (8.6) | 21 (9.1) | 5 (6.8) | 0.551 |
| Rhinobyon or rhinorrhoea | 25 (8.2) | 24 (10.4) | 1 (1.4) | 0.014 |
| Pharyngalgia | 58 (19.1) | 53 (22.9) | 5 (6.8) | 0.002 |
| Dyspnea | 36 (11.8) | 20 (8.7) | 16 (21.9) | 0.002 |
| Diarrhoea | 14 (4.6) | 11 (4.8) | 3 (4.1) | 0.817 |
| Nausea or vomiting | 11 (3.6) | 8 (3.5) | 3 (4.1) | 0.797 |
| **Exposure history** |  |  |  |  |
| Wuhan-related exposure | 136 (44.7) | 94 (40.7) | 42 (57.5) | 0.012 |
| History of close contact | 38 (12.5) | 13 (5.6) | 25 (34.2) | < 0.001 |
| **Days from onset to first hospital admission** | 3.0 (1.0-7.0) | 3.0 (1.0-5.0) | 7.0 (4.0-9.0) | < 0.001 |
| **Basic vital signs** |  |  |  |  |
| <sup>a</sup>Tmax (□) | 37.6 (37.1-38.2) | 37.5 (37.0-38.0) | 38.2 (37.5-39.0) | < 0.001 |
| Body temperature (□) | 37.2 (36.8-37.6) | 37.5 (37.0-37.6) | 37.1 (36.5-37.6) | 0.091 |
| Oxygen saturation (%) | 98.0 (97.0-99.0) | 98.0 (98.0-99.0) | 97.0 (95.0-99.0) | < 0.001 |
| Respiratory (rate breaths/min) | 19.0 (18.0-20.0) | 19.0 (18.0-20.0) | 18.0 (18.0-20.0) | 0.008 |
| Heart rate (beats/min) | 98.0 (86.0-109.0) | 101.50 (89.0-112.0) | 84.0 (76.0-94.0) | < 0.001 |
| Mean arterial pressure (mmHg) | 97.0 (90.1-103.2) | 97.0 (90.7-102.3) | 98.3 (86.7-108.2) | 0.963 |
| **Laboratory results** |  |  |  |  |
| White blood cell count (×10<sup>9</sup>/L) | 6.60 (5.0-8.3) | 6.8 (5.4-8.3) | 5.3 (3.7-8.4) | 0.002 |
| Neutrophil count (×10<sup>9</sup>/L) | 4.1 (2.9-6.2) | 4.1 (3.1-6.1) | 3.7 (2.4-7.0) | 0.356 |
| Lymphocyte count (×10<sup>9</sup>/L) | 1.4 (0.9-1.9) | 1.6 (1.1-2.0) | 0.9 (0.5-1.2) | < 0.001 |
| <sup>a</sup>NLR | 3.0 (1.8-5.8) | 2.7 (1.7-4.7) | 5.0 (2.2-13.9) | < 0.001 |
| Monocyte count (×10<sup>9</sup>/L) | 0.5 (0.4-0.7) | 0.5 (0.4-0.7) | 0.3 (0.2-0.5) | < 0.001 |
| Platelet count (×10<sup>9</sup>/L) | 217.0 (174.5-260.8) | 229.0 (186.0-264.0) | 187.0 (142.0-233.0) | < 0.001 |
| Haemoglobin (g/L) | 139.0 (126.0-153.0) | 139.0 (128.0-155.0) | 137.0 (122.0-150.0) | 0.104 |
| Red blood cell distribution width (%) | 12.3 (11.9-12.8) | 12.3 (12.0-12.8) | 12.20 (11.9-12.8) | 0.227 |
| Platelet distribution width (%) | 11.8 (10.6-13.4) | 11.7 (10.4-13.4) | 12.3 (10.8-13.7) | 0.138 |
| Erythrocyte sedimentation rate (mm/h) | 14.5 (6.0-40.0) | 7.0 (4.0-14.0) | 30.0 (13.5-58.0) | < 0.001 |
| C-reactive protein (mg/L) | 5.3 (1.0-25.0) | 2.1 (1.0-14.4) | 24.3 (9.1-50.5) | < 0.001 |
| Procalcitonin (ng/ml) | 0.15 (0.05-0.24) | 0.20 (0.06-0.25) | 0.07 (0.04-0.10) | < 0.001 |
| Alanine aminotransferase (U/L) | 16.0 (10.0-26.0) | 13.0 (9.0-18.0) | 22.0 (15.0-31.0) | < 0.001 |
| Aspartate aminotransferase (U/L) | 18.0 (15.0-25.0) | 17.0 (14.0-21.2) | 22.0 (16.5-34.5) | < 0.001 |
| Creatinine (μmol/L) | 72.0 (57.3-87.0) | 66.0 (56.0-80.0) | 78.0 (62.5-93.0) | 0.005 |
| Serum urea nitrogen (mmol/L) | 4.6 (3.6-5.8) | 4.3 (3.38-5.0) | 5.2 (3.9-7.0) | 0.002 |
| Lactose dehydrogenase (U/L) | 197.5 (161.0-248.0) | 175.0 (150.5-208.0) | 245.0 (195.0-324.5) | < 0.001 |
| Hydroxybutyrate dehydrogenase (U/L) | 170.0 (139.0-216.0) | 148.0 (130.5-181.5) | 202.0 (168.3-267.5) | < 0.001 |
| **Signs of pneumonia on CT** | 131 (43.1) | 61 (26.4) | 70 (95.9) | < 0.001 |

Data are presented as medians (interquartile ranges, IQR) or N (%).
<sup>a</sup>Tmax: the highest body temperature from illness onset to first hospital admission.

### LABORATORY AND RADIOLOGIC CHARACTERISTICS OF PATIENTS IN THE TRAINING DATASET

As shown in Table 1, the white blood cell count (*P* = 0.002), lymphocyte count (*P* < 0.001), monocyte count (*P* < 0.001), and platelet count (*P* < 0.001) were lower in the COVID-19 group than the non-COVID-19 group. Although neutrophil count had no obvious difference, the neutrophil-to-lymphocyte ratio (NLR) were significantly higher in the COVID-19 group (5.00 [IQR, 2.2-13.9] vs 2.7 [1.7-4.7], *P* < 0.001). Many indicators associated with inflammation or cell damage were increased, including erythrocyte sedimentation rate (ESR), C-reactive protein (CRP), procalcitonin, alanine aminotransferase (ALT), aspartate aminotransferase (AST), lactose dehydrogenase (LDH), and hydroxybutyrate dehydrogenase (HBDH) (all *P* < 0.001). Nearly all patients (95.9%) in the COVID-19 had signs of pneumonia on CT scans. Only 26.4% patients in the non-COVID-19 group had abnormal findings on lung CT scans, and some of them were considered as bacteria or other virus infection.

### DIAGNOSTIC MODEL AND COVID-19 EWS

Variables with *P* < 0.05 were selected to perform multivariate logistic regression analysis by using the forward stepwise method. In order to facilitate the later establishment of a scoring tool, NLR and Tmax were transformed into a categorical variable (< 5.8 / ≥ 5.8; <37.8 / ≥ 37.8) based on the cut-off value. Signs of pneumonia on CT, history of close contact, fever, NLR, Tmax and sex were considered as the independent factors related to the onset of COVID-19 (Supplementary Table 1). The equation was derived: Probability (COVID-19) = 1 / 1 + exp - [−9.106 + (2.79 x Fever) + (4.58 x History of close contact) + (5.10 x Signs of pneumonia on CT) + (0.97 x NLR) + (0.94 x Tmax) + (0.90 x Sex)].

To further improve the operability of this diagnostic model, we converted the beta coefficient values into score. Other significant parameters, age and meaningful lower respiratory symptoms, were also included into the COVID-19 EWS (Figure 2A). According to the results of ROC curve analysis, a cut-off value of 10 was selected as the threshold to distinguish COVID-19 patient, with sensitivity and specificity of 0.932 and 0.874, respectively. As SARS-CoV-2 nucleicacid positive is the accepted standard for COVID-19 diagnosis, and all positive patients need isolated observation and/or treatment, we did not include this single item in COVID-19 EWS.

**Figure 2.**
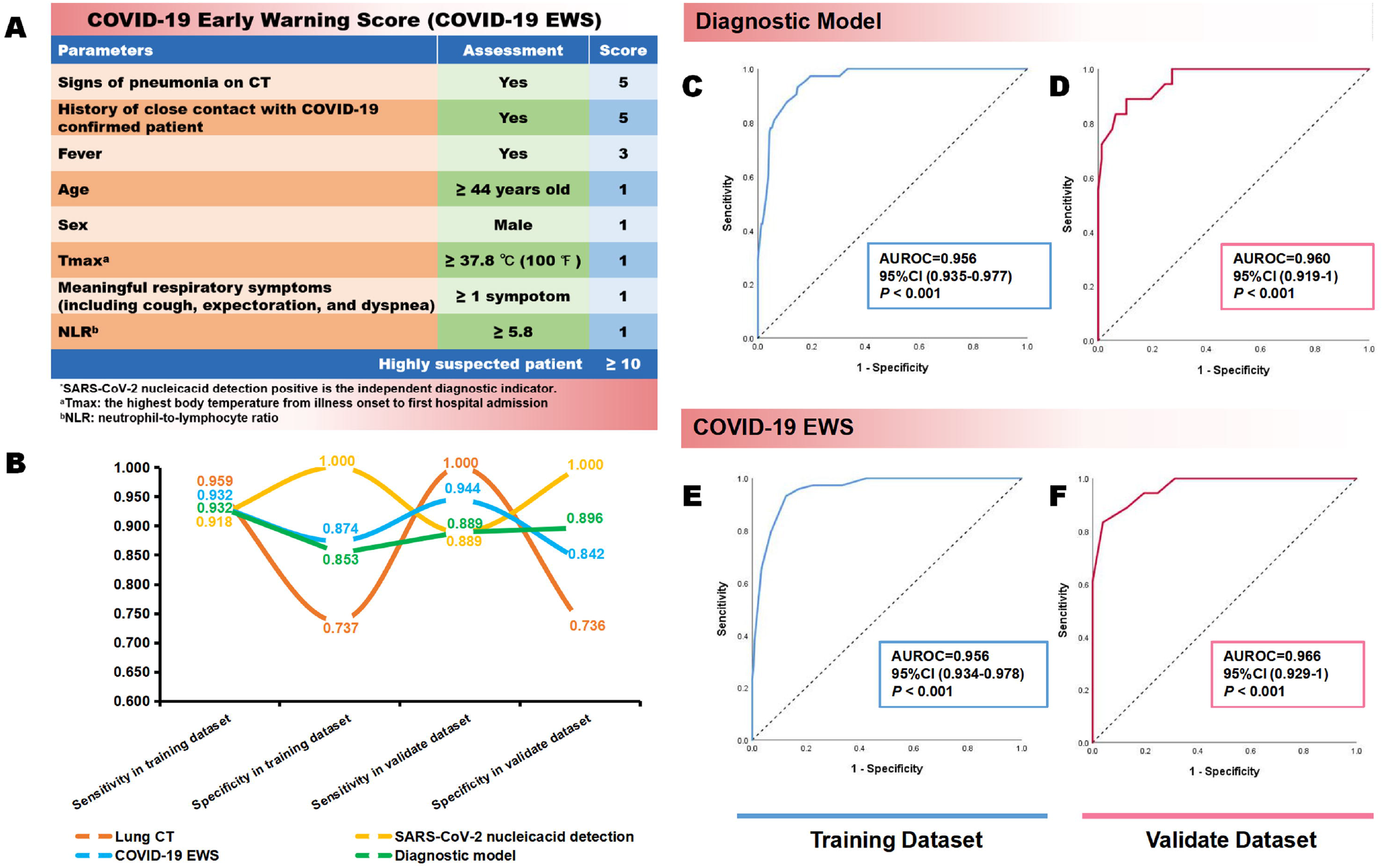
Evaluation of the diagnostic model and COVID-19 Early Warning Score (COVID-19 EWS) COVID-19 EWS was listed (A), patients with a total score of 10 or more were considered highly suspect; the curve graph (B) showed the sensitivity and the specificity of diagnostic model, COVID-19 EWS, SARS-CoV-2 nucleicacid detection and CT scan in training dataset and validate dataset; ROC curves of the diagnostic model and COVID-19 EWS were listed, (C) ROC curve of the diagnostic model in the training group; (D) ROC curve of the diagnostic model in the validation group; (E) ROC curve of the clinical scoring tool in the training group; (F) ROC curve of the clinical scoring tool in the validation group.

In order to determine whether the diagnostic model and COVID-19 EWS are advantageous, we compared their diagnostic capacity to SARS-CoV-2 nucleicacid detection and CT scan (Figure 2B-F). We used the first SARS-CoV-2 nucleicacid detection result to value its sensitivity and specificity. Results showed that although nucleicacid detection had a high specificity, its sensitivity was lower than COVID-19 EWS (training dataset 0.918 vs 0.932; validate dataset 0.889 vs 0.944). Screening only by CT scan had a high sensitivity, but low specificity (training dataset 0.737 vs 0.874; validate dataset 0.737 vs 0.842). Either of the diagnostic model or the COVID-19 EWS had a quite large AUROC both in the training dataset and the validation dataset.

### PREDICTIVE FACTORS ANALYSIS OF COVID-19 DISEASE SEVERITY

We compared the clinical data between the severe group and the non-severe group to find predictive factors of COVID-19 disease severity (Table 2).The median age of the severe group was higher than the non-severe group (55.5 years [IQR 48.0-64.3] vs 48 years [IQR 37.0-59.0], *P* < 0.001), indicating that older patients were more likely to develop severe illness. The proportion of male patients in the severe group was great larger than that of female (71.4% vs 28.6%). Percentage of patients with hypertension was higher in severe group (22 [52.4%] vs 4 [12.9%], *P* < 0.001), while other coexisting disorders, including diabetes, cardiovascular disease and chronic obstructive pulmonary disease showed no significant differences. In addition, many laboratory items and CT score presenting significant differences too. In order to better distinguish severe and non-severe patients, we defined the new threshold value of the selected parameters (*P* < 0.05) by calculating the cut-off value using ROC curve analysis. Table 3 showed the parameters with AUROC > 0.60.

**Table 2.** Comparisons of characteristics between severe patients and non-severe patients with SARS-CoV-2 infection.

| Clinical characteristics | Severe patients<br>(n=42) | Non-severe patients<br>(n=31) | P value |
| --- | --- | --- | --- |
| **Age (years)** | 55.5 (48.0-64.3) | 48.0 (37.0-59.0) | 0.039 |
| **Sex** |  |  | 0.083 |
| Male | 30 (71.4) | 16 (51.6) |  |
| Female | 12 (28.6) | 15 (48.4) |  |
| **Basic vital signs** |  |  |  |
| <sup>a</sup>Tmax (□) | 38.4 (37.5-39.0) | 38.1 (37.5-38.8) | 0.317 |
| Temperature at hospital admission (□) | 37.1 (36.5-39.0) | 37.2 (36.5-37.6) | 0.721 |
| Oxygen saturation (%) | 96.0 (94.5-98.0) | 98.0 (97.0-99.0) | < 0.001 |
| Respiratory (rate breaths/min) | 18.0 (18.0-20.0) | 18.0 (18.0-20.0) | 0.408 |
| Heart rate (beats/min) | 81.5 (69.8-94.0) | 87.0 (78.0-96.0) | 0.126 |
| Mean arterial pressure (mmHg) | 94.0 (85.3-106.4) | 100.0 (91.0-109.3) | 0.418 |
| **Coexisting disorders** |  |  |  |
| Hypertension | 22 (52.4) | 4 (12.9) | < 0.001 |
| Diabetes | 4 (9.5) | 2 (6.5) | 0.637 |
| Cardiovascular disease | 4 (9.5) | 1 (3.2) | 0.292 |
| Chronic obstructive pulmonary disease | 2 (4.8) | 1 (3.2) | 0.744 |
| Chronic liver disease | 1 (2.4) | 3 (9.7) | 0.176 |
| **Laboratory results** |  |  |  |
| White blood cell count (×10<sup>9</sup>/L) | 6.6 (3.9-11.0) | 4.2 (3.5-6.0) | 0.007 |
| Neutrophil count (×10<sup>9</sup>/L) | 5.8 (2.8-10.2) | 2.8 (1.8-4.1) | 0.002 |
| Lymphocyte count (×10<sup>9</sup>/L) | 0.8 (0.5-1.1) | 1.0 (0.6-1.3) | 0.085 |
| <sup>a</sup>NLR | 8.2 (3.9-19.2) | 3.0 (1.9-5.5) | 0.001 |
| Monocyte count (×10<sup>9</sup>/L) | 0.3 (0.2-0.5) | 0.4 (0.3-0.5) | 0.163 |
| Platelet count (×10<sup>9</sup>/L) | 189.0 (154.0-231.0) | 178.0 (127.0-239.0) | 0.321 |
| Haemoglobin (g/L) | 136.5 (120.8-152.0) | 137.0 (126.0-149.0) | 0.920 |
| Red blood cell distribution width (%) | 12.4 (12.1-13.0) | 11.9 (11.8-12.6) | 0.007 |
| Platelet distribution width (%) | 12.3 (11.0-13.5) | 12.8 (10.7-13.9) | 0.643 |
| Erythrocyte sedimentation rate (mm/h) | 46.0 (21.3-68.5) | 20.0 (10.0-40.0) | 0.016 |
| C-reactive protein (mg/L) | 35.0 (16.1-59.6) | 10.3 (3.5-30.0) | 0.001 |
| Procalcitonin (ng/ml) | 0.07 (0.04-0.10) | 0.06 (0.04-0.10) | 0.267 |
| Alanine aminotransferase (U/L) | 21.0 (13.0-30.0) | 25.0 (16.0-36.8) | 0.350 |
| Aspartate aminotransferase (U/L) | 22.0 (16.5-41.0) | 21.0 (16.5-29.8) | 0.673 |
| Creatinine (μmol/L) | 82.0 (67.0-102.0) | 68.0 (56.3-88.8) | 0.057 |
| Serum urea nitrogen (mmol/L) | 5.6 (4.6-8.5) | 4.4 (3.4-5.9) | 0.012 |
| Lactose dehydrogenase (U/L) | 287.0 (220.0-349.5) | 223.0 (185.0-258.8) | 0.003 |
| Hydroxybutyrate dehydrogenase (U/L) | 233.0 (178.3-302.3) | 181.0 (151.0-216.0) | 0.002 |
| Total T cell count (10<sup>6</sup>/L) | 269.0 (158.0-410.0) | 504.5 (262.0-918.8) | 0.025 |
| CD4<sup>+</sup>T cell count (10<sup>6</sup>/L) | 139.0 (72.0-206.0) | 288.5 (142.5-504.0) | 0.011 |
| CD8<sup>+</sup>T cell count (10<sup>6</sup>/L) | 117.0 (59.0-177.0) | 234.0 (122.3-367.8) | 0.005 |
| CD4/CD8 | 1.2 (1.1-1.7) | 1.2 (0.9-1.6) | 0.790 |
| NK cell count (10<sup>6</sup>/L) | 100.0 (54.0-178.0) | 232.5 (100.5-286.3) | 0.013 |
| B cell count (10<sup>6</sup>/L) | 82.0 (45.0-149.0) | 115.5 (83.0-161.5) | 0.203 |
| IL-2 (pg/ml) | 1.0 (0.8-1.9) | 1.0 (0.7-2.0) | 0.956 |
| IL-4 (pg/ml) | 1.8 (1.4-1.8) | 1.8 (1.2-1.8) | 0.437 |
| IL-6 (pg/ml) | 24.2 (11.6-47.0) | 21.6 (8.7-57.2) | 0.869 |
| IL-10 (pg/ml) | 6.7 (3.3-8.2) | 4.3 (3.0-8.1) | 0.406 |
| TNF-α (pg/ml) | 12.2 (12.2-54.4) | 19.7 (2.5-67.5) | 0.581 |
| IFN-γ (pg/ml) | 9.0 (5.7-24.3) | 12.2 (5.8-37.9) | 0.452 |
| **CT score** | 15.0 (11.5-18.3) | 6.0 (3.0-8.0) | < 0.001 |

Data are presented as medians (interquartile ranges, IQR) or N (%).
<sup>a</sup>NLR: neutrophil-to-lymphocyte ratio

**Table 3.** Cut-off values of predictive factors analysis of COVID-19 disease severity

| Parameters | AUROC | Cut-off Value | 95% CI | Sensitivity | Specificity |
| --- | --- | --- | --- | --- | --- |
| CT score | 0.98 | 8.5 | 0.95-1.00 | 0.95 | 0.94 |
| CD8<sup>+</sup> T cell count (10<sup>6</sup>/L) | 0.76 | 191.0 | 0.62-0.90 | 0.64 | 0.80 |
| CD4<sup>+</sup> T cell count (10<sup>6</sup>/L) | 0.74 | 211.5 | 0.58-0.89 | 0.71 | 0.80 |
| Age (years) | 0.74 | 44 | 0.68-0.80 | 0.75 | 0.70 |
| NK cell count (10<sup>6</sup>/L) | 0.73 | 214.0 | 0.56-0.90 | 0.57 | 0.89 |
| C-reactive protein (mg/L) | 0.73 | 14.22 | 0.60-0.85 | 0.83 | 0.58 |
| <sup>a</sup>NLR | 0.72 | 5.87 | 0.60-0.84 | 0.64 | 0.81 |
| Hydroxybutyrate dehydrogenase (U/L) | 0.72 | 200.50 | 0.60-0.84 | 0.70 | 0.71 |
| Total T cell count (10<sup>6</sup>/L) | 0.71 | 455.5 | 0.54-0.88 | 0.71 | 0.80 |
| Serum urea nitrogen (mmol/L) | 0.68 | 4.54 | 0.55-0.81 | 0.78 | 0.57 |
| Erythrocyte sedimentation rate (mm/h) | 0.67 | 26.00 | 0.54-0.79 | 0.74 | 0.61 |

<sup>a</sup>NLR: neutrophil-to-lymphocyte ratio

## DISCUSSION

The worldwide outbreak of COVID-19 is intensifying.^12^ As the main battlefield on the first stage, early detection and effective quarantine of patients and close contacts has made the epidemic in China so far under effective control. However, practice has shown that detection of highly suspected patients remains problematic. Considering the relative shortage of SARS-CoV-2 nucleicacid detection kits and false-negative results caused by various reasons, such as the quality of the samples taken, the number of viruses and the stage of the disease, medical experts have proposed screening of highly suspected patients with lung CT. However, due to mild COVID-19 patients are defined as no pneumonia on imaging, and imaging findings of some patients are atypical, screening based on CT findings is greatly dependent on the physician’s experience and the effectiveness is limited. All the above reasons increase the diagnosis difficult of COVID-19 patients when they first come to the doctors.

In our study, we firstly developed a diagnostic model based on multivariate logistic regression analysis. Since this diagnostic model was a logistic regression equation and inconvenient to operate in clinical environment, we converted the diagnostic model into an scoring tool by assigning values to each variable in the equation. Besides of that, we enrolled two more parameters into the final COVID-19 EWS, including age and meaningful respiratory symptoms. Based on the statistical analysis, we found that elderly people were more susceptible to COVID-19 and develop severe, which may due to their lower disease resistance and more basic diseases; some significant symptoms, such as cough, expectoration and dyspnea, can help to strengthen the association between fever and lower respiratory symptoms and weaken the weight of fever caused by other reasons. As a result, the COVID-19 EWS contained easy-to-get parameters on clinical manifestation, epidemiological characteristics, basic vital signs, laboratory and radiologic data, which provide a more comprehensive assessment of patients. Through re-evaluation in the validation dataset and comparison with other methods, results showed that both of our diagnostic model and COVID-19 EWS had relatively high and stable sensitivity and specificity.

The clinical features of patients in our medical center were similar to those reported in previous studies^13-15^: Male were more susceptible to COVID-19 but had no significance to predict of disease severity in our study may due to the small sample size, although the male to female ratio was quite high in the severe group (71.4 vs 28.6); most patients had initial symptoms of fever and cough; as for laboratory results, the decrease of white blood cell, lymphocyte, monocyte, and platelet, and the increase of the levels of ESR, CRP, Cr, serum urea nitrogen, LDH, and HBDH were found in COVID-19 patients, suggesting that SARS-CoV-2 infection may be associated with the injury of cellular immune, cardiomyocytes, liver and kidney.^16^ NLR was a widely used marker for the assessment of the severity of bacterial infections and the prognosis of patients with pneumonia and tumors.^17-19^ In our study, we also found the increase of NLR in COVID-19 patients, and it was positively related to the disease severity.

Consistent with a previous study, we found CD4^+^ T cell count and CD8^+^ T cell count were negatively correlated to the severity of COVID-19, indicating that SARS-CoV-2 may direct or indirect damage to T lymphocytes and further aggravates disease progression.^20,21^ Although inflammatory cytokine storms were thought to be a mechanism for COVID-19 progression,^15^ we did not find an significantly increased level of IL-2, IL-4, IL-6, IL-10, TNF-α, and IFN-γ. These findings might due to the limited size of our study and the large heterogeneity at the time-points of the first detection.

There were also many shortcomings in our study that should be considered. Firstly, it was a retrospective study, which might contain selection bias, although we tried to avoid the bias through sorting patients by “Z-A” based on Chinese first name and strictly abide the inclusion and exclusion criteria. In addition, because of the incomplete of potentially valuable information, such as smoking history and basic diseases, we cannot conduct a comprehensive analysis of these factors currently. Besides of that, additional multi-center, multi-ethnic and prospective studies are expected to revised our diagnostic model and COVID-19 EWS, and we also plan to implement a multi-center study with a larger sample size to further validate and optimize the model. More over, it is hoped that better statistical algorithms will make the diagnostic model more practical.

## CONCLUSION

We established a novel and easy-to-get early warning score for COVID-19 screening. This scoring tool allows clinicians to more quickly and relatively accurately detect COVID-19 patients, especially when the nucleicacid detection capacity is relatively lacking.

## Data Availability

The data used in this study are available from the corresponding author.

## ACKNOWLEDGMENTS

We thank for Ying-ying Mao, Ph.D., providing the help on statistical analyses.

